# Gestational Weight Gain Management in Underserved Mothers - A State-Wide Randomized Controlled Trial in Louisiana WIC

**DOI:** 10.1101/2025.01.29.25321347

**Authors:** Emily W. Flanagan, Kaja Falkenhain, Robbie Beyl, Abby D. Altazan, S. Ariel Richard, Hannah E. Cabre, Chelsea L. Kracht, Joshua R. Sparks, Maryam Kebbe, L. Anne Gilmore, Daniel S. Hsia, John W. Apolzan, Leanne M. Redman

## Abstract

**Importance:** Underserved pregnant individuals experience the highest risk of aberrant pregnancy weight gain and adverse perinatal outcomes. The Women, Infants, and Children (WIC) federal program assists underserved pregnant individuals and is therefore positioned to offer equitable access to interventions to enhance gestational weight gain in accordance with clinical guidelines.

**Objective:** Test effectiveness of a pragmatic, fully remote lifestyle intervention co-developed with WIC participants on the incidence of gestational weight gain guideline attainment and perinatal outcomes.

**Design:** **The** SmartMoms in WIC trial was a single-blind randomized clinical trial conducted from July 2019 to May 2024.

**Setting:** Louisiana WIC Program pregnant participants across 31 participating WIC clinics.

**Participants:** 1300 individuals were recruited from Louisiana WIC; 756 were excluded via phone call and 544 were screened in person; 351 were enrolled. Randomization was stratified by geographical region and BMI class.

**Intervention:** A high intensity multicomponent e-health intervention (“Healthy Beginnings”) for gestational weight gain management or usual care between 10 to 16 weeks gestation and until delivery (∼24 weeks).

**Main Outcome(s) and Measure(s):** The primary outcome was assessed at participants’ WIC clinic and included gestational weight gain guideline attainment with total observed weight gain, weight gain per week and deviation from guidelines as secondary outcomes. Perinatal outcomes from birth certificates were exploratory.

**Results:** The study sample (179 Intervention; 172 Usual Care) was diverse: 39% with obesity; 57% non-Hispanic Black. The incidence of guideline attainment was not different between groups. Study observed total (adjusted mean difference, –1.4 kg; 95% CI, –2.8 to –0.1), and rate of weight gain (adjusted mean difference –0.07 kg/wk; 95% CI, –0.13 to –0.01) and the deviation from guidelines was lower in the Intervention Group compared to Usual Care. There were 43 cases (16/172 Intervention, 27/171 Usual Care) of preterm birth and 30 NICU admissions (12/172 Intervention, 18/171 Usual Care) equating to an adjusted relative risk reduction of 36.9% and 28.6%, respectively.

**Conclusions and Relevance:** A fully remote lifestyle intervention concomitant with WIC clinical care lowered gestational weight gain and reduced the risk of preterm birth and NICU admission.

**Trial Registration:** ClinicalTrials.gov NCT04028843

**ABSTRACT:** *Background:* Underserved pregnant individuals experience the highest risk for weight gain outside clinical guidelines and adverse perinatal outcomes. The Women, Infants, and Children (WIC) federal program assists underserved pregnant individuals with supplemental nutrition. WIC is positioned to offer equitable access to interventions promoting recommended gestational weight gain, complementing clinical care.

*Methods:* In a state-wide randomized controlled trial, pregnant WIC participants were randomly assigned to a co-developed multicomponent e-health intervention for gestational weight gain management or usual care between 10 to 16 weeks gestation. The primary outcome was gestational weight gain guideline attainment. Study observed weight gain, weight gain per week and deviation from guidelines were secondary outcomes. Perinatal outcomes from birth certificates were exploratory.

*Results:* Pregnant participants (n=351) were enrolled (179 Intervention; 172 Usual Care) across 31 WIC clinics. The study sample was diverse: 39% with obesity; 57% non-Hispanic Black. The incidence of guideline attainment was not different between groups. Study observed weight gain (adjusted mean difference, –1.4 kg; 95% CI, –2.8 to –0.1), rate (adjusted mean difference, –0.07 kg/wk; 95% CI, –0.13 to –0.01) and the deviation from guidelines was lower in the Intervention Group compared to Usual Care. There were 43 cases (16/172 Intervention, 27/171 Usual Care) of preterm birth and 30 NICU admissions (12/172 Intervention, 18/171 Usual Care) equating to an adjusted relative risk reduction of 36.9% and 28.6%, respectively.

*Conclusion:* A fully remote lifestyle intervention concomitant with WIC clinical care lowered gestational weight gain and reduced the risk of preterm birth and NICU admission.

*Trial registration:* ClinicalTrials.gov NCT04028843

**Key Points:** *Question:* Can a pragmatic, fully remote lifestyle intervention improve gestational weight gain and perinatal outcomes in underserved pregnant WIC participants?

*Findings:* In this state-wide randomized controlled trial that included 351 pregnant WIC participants, the incidence of National Academy of Medicine guideline attainment for weight gain did not differ. Study observed gestational weight gain, preterm birth, and NICU admissions were lower in the Intervention Group.

*Meaning:* A fully remote lifestyle intervention concomitant with WIC clinical care attenuated gestational weight gain and improved perinatal outcomes.

## INTRODUCTION

MATERNAL MORBIDITY IN THE UNITED STATES is one of the highest among developed countries, with underrepresented and underserved communities disproportionately affected. The risk for adverse perinatal conditions is increased by aberrant gestational weight gain which is experienced by three out of four pregnant people^1,2^. There is cross-cutting agreement that weight during pregnancy be managed with prenatal lifestyle programs in primary care and public health settings according to recommended guidelines^3,4^.

Towards the implementation of a pragmatic and patient-centered program, we partnered with stakeholders to co-adapt an evidence-based multi-component (i.e., dietary modification, physical activity modification, behavioral counselling) lifestyle intervention to be fully remote, of appropriate health literacy, and tailored to pregnant people enrolled in the Louisiana Special Supplemental Nutrition Assistance, Women, Infants, and Children (WIC) program^5^. WIC serves 6.7 million mothers and children annually^6^, and therefore has an unparalleled ability to reach economically disadvantaged and health disparate communities. This reports the primary outcome of “SmartMoms in WIC”, a state-wide randomized controlled trial embedded in the Louisiana WIC program^7^, to test the effectiveness of the cooperatively developed pragmatic intervention to promote recommended gestational weight gain and determine effects on perinatal outcomes.

## METHODS

### STUDY DESIGN

This trial was a parallel arm randomized controlled trial conducted by an academic research team embedded within the Louisiana WIC program. The trial protocol is available in **Supplemental Appendix** and has been published previously^7^. Participants provided informed consent for study procedures approved by the institutional review boards of Pennington Biomedical Research Center and the Louisiana Department of Health.

The trial was designed to test the hypothesis that participants receiving usual WIC Nutrition and the Healthy Beginnings intervention (Intervention Group) will have a higher incidence of appropriate weight gain according to the 2009 National Academy of Medicine (NAM) gestational weight gain per week guidelines^8^ compared to participants receiving usual WIC Nutrition only (Usual Care Group). Secondary outcomes included study observed gestational weight gain (total and per week) and deviation in gestational weight gain per week from guidelines.

Subgroup analyses were planned a-priori for intervention effects within BMI categories (normal weight, overweight, obesity). Perinatal outcomes abstracted from birth certificates were exploratory.

### PARTICIPANTS

Recruitment occurred between July 2019 and December 2023 within 31 participating WIC clinics across Louisiana which were selected based on size and location. Pregnant individuals certified to receive the WIC pregnancy benefit package were introduced to the trial at their WIC clinic, or via a telephone call or text message by study staff. Eligible participants were 18 to 40 years, pregnant with a singleton gestation, with a body mass index (BMI) between 18.5 and 40.0 kg/m^2^ measured at screening (<16 weeks gestation), owned a smartphone with internet access, and were willing to be identifiable to other participants in private social media groups. Exclusion criteria were reported previously and are shown in **Fig. 1**.

**Figure 1.**
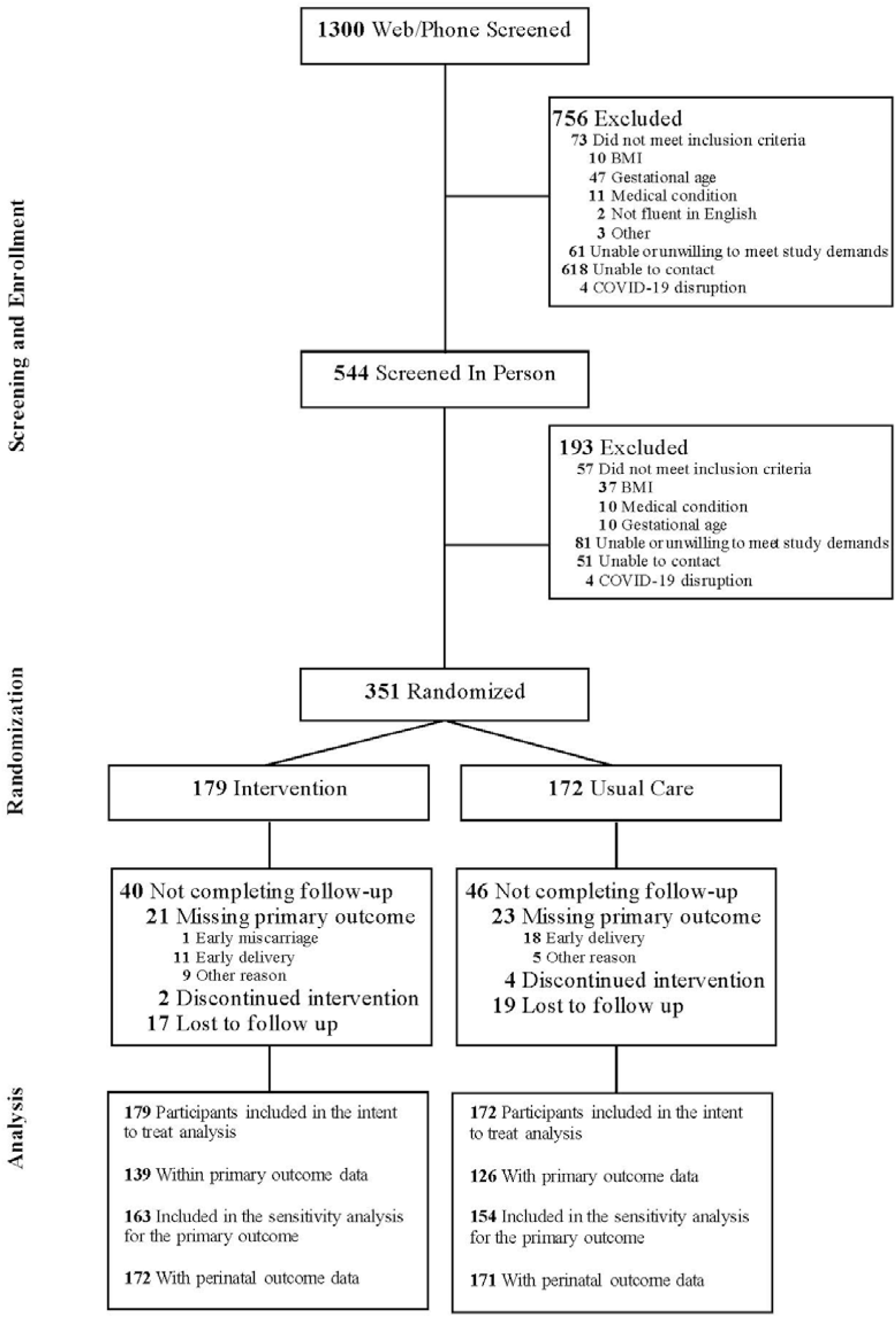
Screening, Enrollment, Randomization, and Inclusion in Analysis. A total of 351 participants were included in the intention-to-treat analysis, of which 265 (75%) had weight obtained at the end of pregnancy. The sensitivity analysis for the primary outcome included participants with weight at mid-pregnancy assessment when end of pregnancy assessment was missing. All participants with birth certificate data were included in the perinatal outcomes analysis. Early delivery is defined as delivery prior to scheduled outcome visit.

### TRIAL PROCEDURES

Participants were randomized 1:1 to the Healthy Beginnings (Intervention Group) or WIC Nutrition (Usual Care Group) after baseline assessments (**Fig. 1**). Randomization used a permuted block strategy with stratification for clinic geographical region (1 through 9, as defined by the Louisiana Department of Health) and BMI class (normal weight, overweight, obesity), prepared by the trial statistician. The Research Electronic Data Capture (REDCap) platform was used to conceal allocation sequence to intervention staff prior to group assignment. The trial adhered to a single-blind design with outcome assessors and interventionists kept separate.

### TRIAL INTERVENTIONS

#### Usual Care Group

The Usual Care Group received all aspects of the Louisiana WIC program, a brief monthly call to bond them with the trial and access to a private Facebook group where interventionists shared weekly posts unrelated to weight.

#### Intervention Group

The Healthy Beginnings intervention was co-developed with the Baton Rouge Community Advisory Board of the Louisiana Clinical and Translational Science Center and a WIC Mothers’ Advisory Group established for the trial^7^. The intervention was adapted from SmartMoms^5^ and was a ∼24-week, evidence-based multicomponent behavioral modification program to coach individuals on achieving BMI-appropriate gestational weight gain^7^. The behavioral intervention was delivered via weekly lessons via two-to four-minute videos and an accompanying transcript of health information and behavior change strategies. The videos were presented by intervention staff alongside actors who were pregnant and with likeness to the trial participants. Lessons were reinforced during weekly virtual check-ins with health coaches and additional cooking and exercise videos were posted in a private Facebook group. Participants were prescribed self-monitoring of weight and physical activity (steps per day) and received automated prescriptive feedback on their weight change. To encourage adherence, participants were incentivized with a gamification program. Points earned for self-monitoring behaviors^5^, coach check-ins and lesson viewing could be redeemed each month for items associated with postpartum and newborn care. All interactions and treatment recommendations between the participant and coaches occurred via the multi-media functions of the smartphone. Coaches with college degrees in nutrition, kinesiology or psychology followed a treatment manual and were supervised by one of the trial investigators. Intervention fidelity was guided by the treatment manual and weekly case conferencing.

Intervention participants were mailed a toolkit which included a cellular-enabled electronic scale and an activity monitor with instructions to synchronize the devices with their phone and to access the Healthy Beginnings online portal. Participants had a 20-minute telephone call with their coach to review the goals for the intervention and orientation with the equipment, participant website, gamification program and to establish weekly check-ins.

Self Monitoring Component: The cornerstone of the intervention was a NAM weight gain chart personalized to each participant beginning with their weight at randomization and BMI-specific weight gain per week goals. Participants were encouraged to weigh daily but at least weekly and their weight gain trajectory prompted automated feedback.

Dietary Component: Diet-based lessons encouraged adherence to the US Dietary Guidelines and utilization of the WIC food package with co-developed meal plans, shopping lists, recipes and cooking demonstration videos. Participants also received measuring cups and spoons, mixing bowls and a reusable drink bottle and lunch box.

Physical Activity Component: The intervention encouraged 150 minutes per week of physical activity, and participants set goals for daily steps which were monitored by the Fitbit and tracked on a step chart. Participants had access to eight exercise videos, received instructions for a new exercise each week and were provided with resistance bands, a pair of 2 lb dumbbells and an exercise mat.

### OUTCOMES

Weight was measured in duplicate using a standardized calibrated electronic scale (Tanita Corp, Arlington Heights, IL) to the nearest 0.1 kg with the participant wearing light clothing and assessed by study staff prior to randomization in early pregnancy (10,0 to 16,6 weeks gestation), mid-pregnancy (24,0 to 27,6 weeks) and late pregnancy (35,0 to 37,6 weeks). The primary outcome was the incidence of appropriate gestational weight gain per week observed between early and late pregnancy^8^. Secondary outcomes included study observed total (kg) and rate of (kg/wk) gestational weight gain, and the deviation in weight (kg) from recommended guidelines. Incidence of perinatal outcomes (Maternal: gestational diabetes; hypertension/preeclampsia; medically indicated Cesarean section; delivery prior to 37 weeks; Neonatal: small for gestational age [weight <10^th^ percentile]; large for gestational age [weight >90^th^ percentile]; neonatal intensive care unit [NICU] admission) abstracted from birth certificates were exploratory. Engagement with the intervention was evaluated using four metrics; responsiveness to coach contacts, video lesson viewing and the frequency of self-monitoring of body weight and steps. Detailed definitions of study outcomes are provided in **Supplemental Appendix**.

### STATISTICAL ANALYSIS

The trial was powered based on the Louisiana Pregnancy Risk Assessment Monitoring System and our pilot trial^5^ to detect a difference in the incidence of appropriate weight gain of 14% between groups with an overall power of 80% and a two-sided type 1 error rate of less than 5%. Inflating by 15% to account for typically observed attrition rates^10^, the initially targeted sample size was 432. The COVID-19 pandemic caused a five-month recruitment halt. To conclude enrollment by December 2023, the trial investigators adjusted sample size estimates to detect a difference of 16% in the overall incidence of appropriate weight gain between groups with 80% power. The revised sample size required a minimum of 330 participants, while retaining adequate power to detect intervention effects on secondary outcomes as well as within BMI categories (statistical analysis plan presented in **Supplemental Appendix**).

Blinded data were analyzed on an intention-to-treat basis. Data are presented as mean (SD) or N (%) for continuous and categorical variables, respectively. The primary outcome of incidence of gestational weight gain guideline attainment was analyzed using a linear model with a binary distribution and a log odd link function. Secondary outcomes of total gestational weight gain was analyzed using a linear mixed-effects model including fixed effects for time (early pregnancy, late pregnancy), group (Intervention, Usual Care), and its interaction, with participant as a random effect to account for within-subject repeated measures. Secondary outcomes of weekly gestational weight gain and deviation from recommended weight gain ranges were analyzed using a linear model with a fixed effect for group. All models included parity (nulliparous, non-nulliparous), BMI category (normal weight, overweight, obesity) at randomization and gestational age at randomization as covariates. The prespecified subgroup analysis of intervention effects within each BMI category was tested using custom comparisons with a three-way interaction (time, group, and BMI category) and adjustments for the same covariates. Perinatal outcomes were additionally analyzed using a linear model with a binary distribution and a log odd link function modeling the incidence. Model-adjusted mean differences between groups or adjusted odds ratios alongside 95% confidence interval and associated P value are reported as the main effects of interest. All analyses were conducted with the use of R, version 4.3.2 (R Core Team)^11^.

## RESULTS

### PARTICIPANT CHARACTERISTICS

A total of 351 pregnant individuals were enrolled with 179 assigned to the Intervention Group and 172 to the Usual Care Group (**Fig.1**). Mean gestational age at randomization was 15 weeks, 1 day. The sample included 114 (33%) participants with normal weight, 99 (28%) overweight, and 138 (39%) with obesity. The majority of participants identified as non-Hispanic Black (57%). Overall, characteristics of the Intervention and Usual Care Groups were comparable at enrollment (**Table 1**). Clinic measured weight at the end of pregnancy was available for 265 (75%) participants. Missing data was due to 30 miscarriage or early delivery (9%), 14 (4%) skipped visit; 6 (2%) discontinued intervention; and 36 (10%) lost to follow up.

**Table 1.** Baseline Characteristics of Study Participants.□*.

| Characteristic | Total<br>(N=351) | Intervention<br>(N=179) | Usual Care<br>(N=172) |
| --- | --- | --- | --- |
| **Gestational age at randomization – weeks** | 15.2±1.6 | 15.2±1.6 | 15.1±1.6 |
| **Maternal age – years** | 27±6 | 27±6 | 28±6 |
| **Nulliparous – no. (%)** | 150 (43) | 84 (47) | 66 (38) |
| **Body mass – kg** | 75.1±16.9 | 74.8±17.5 | 75.4±16.4 |
| **Body mass index <sup>†</sup>** | 28.5 ± 5.8 | 28.6±5.9 | 28.5±5.7 |
| □□□□ Normal weight; 18.5-24.9 – no. (%)□ | 114 (33) | 58 (32) | 56 (33) |
| □□□□ Overweight; 25.0-29.9 – no. (%)□ | 99 (28) | 51 (29) | 48 (28) |
| □□□□ Obesity; 30.0-40.0 – no. (%)□ | 138 (39) | 70 (39) | 68 (39) |
| **Race/Ethnicity – no. (%)□** |  |  |  |
| □□□□ Hispanic□ | 23 (7) | 10 (6) | 13 (8) |
| □□□□ Non-Hispanic White□ | 109 (31) | 55 (31) | 54 (31) |
| □□□□ Non-Hispanic Black□ | 201 (57) | 106 (59) | 95 (55) |
| □□□□ Mixed or Other□ | 18 (5) | 8 (4) | 10 (6) |
| **Marital status – no. (%)□** |  |  |  |
| □□□□ Married/Living with significant other□ | 200 (57) | 104 (58) | 96 (56) |
| □□□□ Not married□ | 151 (43) | 75 (42) | 76 (44) |
| **Educational attainment – no. (%) □** |  |  |  |
| □□□□ Some college education□□□□□ | 221 (63) | 110 (61) | 111 (65) |
| □□□□ No college education□ | 130 (37) | 69 (39) | 61 (35) |

\* Plus-minus values are means ± SD. <sup>†</sup>Body mass index is the weight in kilograms divided by height in meters squared.

State abstracted birth certificate data were available for 343 (98%) participants. Participant demographics of those not assessed at end of pregnancy were similar to those assessed (**eTable 1**).

### GUIDELINE ATTAINMENT

The incidence of appropriate gestational weight gain according to NAM guidelines as determined by the rate of weight gain per week was similar between the Intervention and Usual Care Groups at the end of pregnancy (both groups 17%; adjusted odds ratio, 1.05, 95% confidence interval [CI], 0.55 to 2.01; P=0.89) (**Table 2**).

**Table 2.**
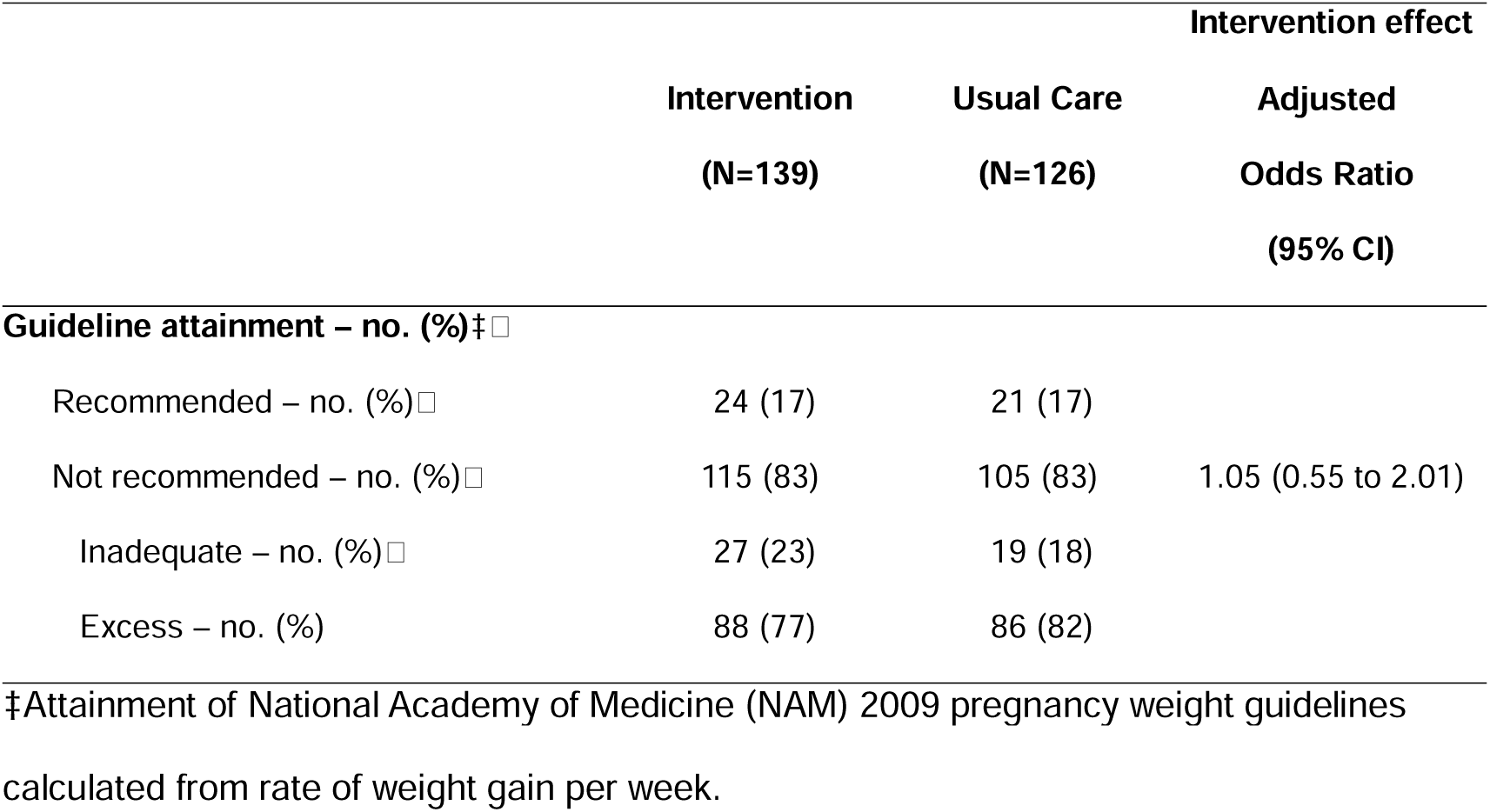
Gestational Weight Gain Guideline Attainment.

### GESTATIONAL WEIGHT GAIN

Study observed weight gain was lower in the Intervention Group (9.5 kg; 95% CI, 8.5 to 10.5 kg) compared to the Usual Care Group (10.9 kg; 95% CI, 9.9 to 11.9 kg) across the intervention period (adjusted mean difference, –1.4 kg; 95% CI, –2.8 to –0.1 kg; P=0.04; **Fig. 2A**). Rate of weight gain was lower in the Intervention Group (0.46 kg; 95% CI, 0.42 to 0.50 kg) compared to the Usual Care Group (0.53 kg; 95% CI, 0.48 to 0.57 kg) with an adjusted mean difference of –0.07 kg (95% CI, –0.13 to –0.01 kg) between groups (P=0.02; **Fig. 2B**). The deviation in weight gain per week from NAM guidelines was lower in the Intervention Group (0.19 kg; 95% CI, 0.16 to 0.23 kg) compared to the Usual Care Group (0.25 kg; 95% CI, 0.21 to 0.28 kg) with an adjusted mean difference of –0.05 kg (95% CI, –0.10 to –0.00 kg; P=0.03; **Fig. 2C**). Sensitivity analyses including weight data recorded at the mid-pregnancy visit for those with missing data at the end of pregnancy showed similar results (**eTable 2A-B**).

**Figure 2.**
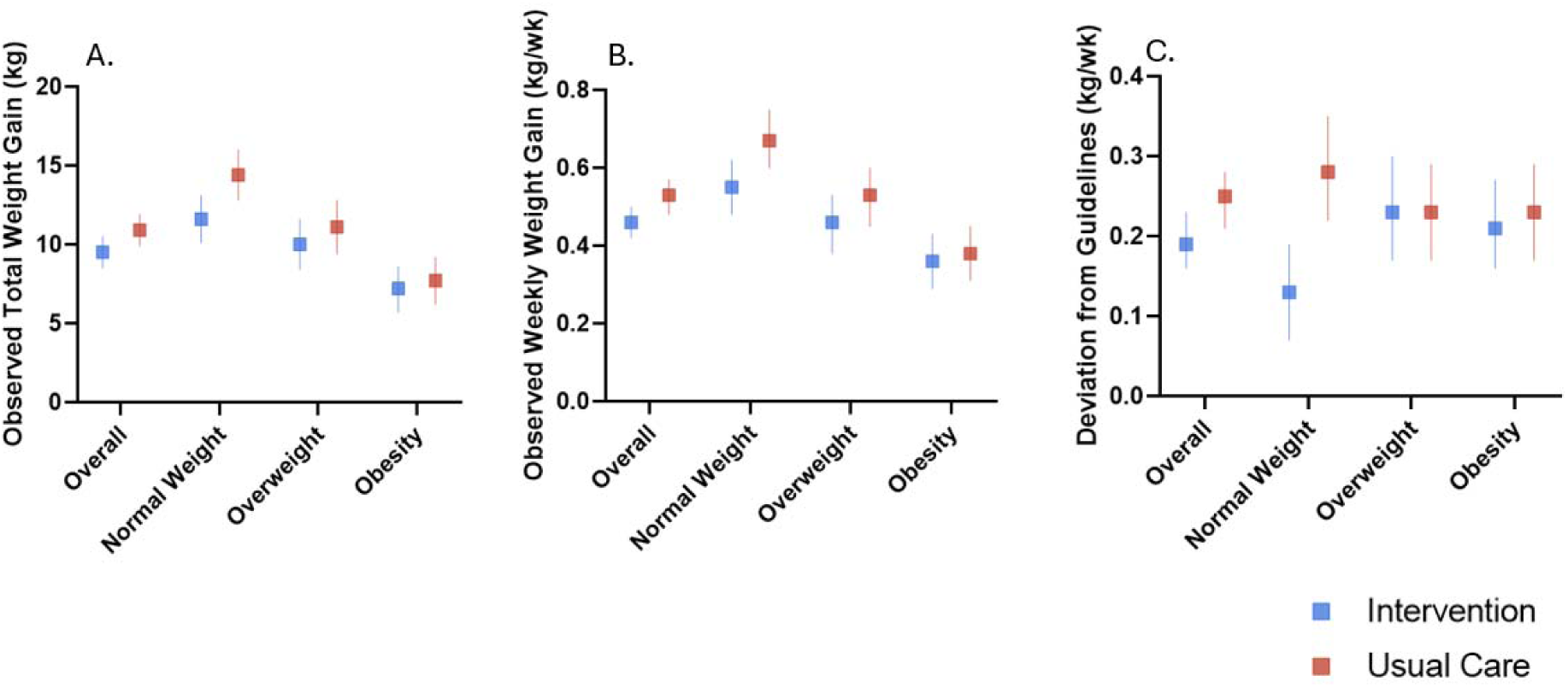
Gestational Weight Gain. Effect estimates and 95% confidence intervals presented for (A) study observed total weight gain; (B) rate of weight gain; and (C) deviation from National Academies of Medicine guidelines.

### BMI SUBGROUPS

Guideline attainment in the normal weight stratum and all gestational weight gain outcomes in the overweight and obesity strata were similar between Intervention and Usual Care Groups **(eTable 3A-F).** Weight gain outcomes in the normal weight group demonstrated a significant intervention effect for study observed gestational weight gain (adjusted mean difference, –2.8 kg; 95% CI, –5.0 to –0.6 kg; P=0.01; **Fig. 2A**), rate of gestational weight gain (adjusted mean difference, –0.12 kg; 95% CI: –0.22 to –0.02 kg; P=0.02; **Fig. 2B**) and deviation in gestational weight gain from guidelines (adjusted mean difference, –0.15 kg; 95% CI, –0.24 to –0.06 kg; P=0.001; **Fig. 2C**).

### PERINATAL OUTCOMES

Adjusted odds ratios of maternal outcomes are shown in **Fig. 3A**. The birth certificates report 26 cases of gestational diabetes (13 Intervention, 13 Usual Care), 55 cases of gestational hypertension (26 Intervention, 29 Usual Care), 73 cases of medically indicated Cesarean (36 Intervention, 37 Usual Care), and 43 cases of preterm delivery prior to 37 weeks (16 Intervention, 27 Usual Care). The incidence of preterm delivery was lower in the Intervention Group (9% vs 16% in Usual Care; adjusted OR, 0.56; 95% CI, 0.28 to 1.08), which equated to an adjusted relative risk reduction of 36.9%. There were 6 reports of maternal morbidity (3 Intervention, 3 Usual Care), which composed of: maternal transfusion (3), third-or fourth-degree perineal laceration (3), admission to the intensive care unit (1) and unplanned operative procedure following delivery (1).

**Figure 3.**
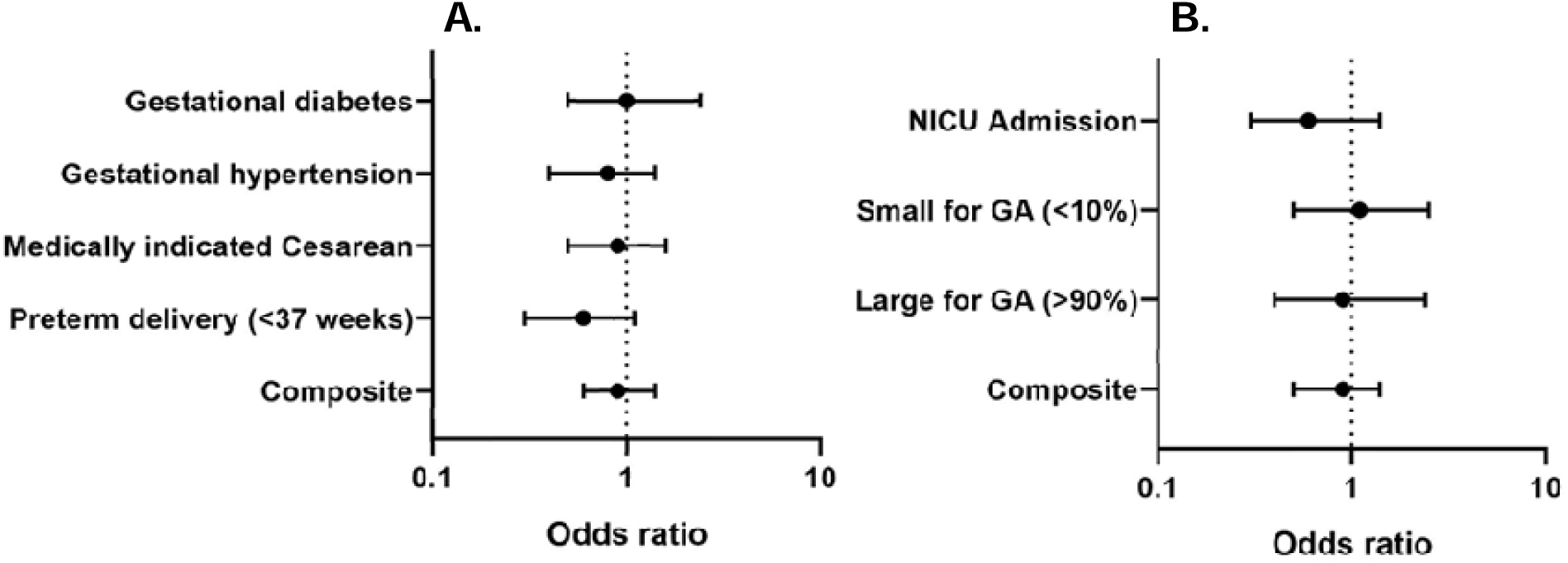
Perinatal Outcomes. Adjusted odds ratios presented for (A) maternal outcomes and (B) neonatal outcomes. NICU denotes neonatal intensive care unit; GA denotes gestational age. Composite is a binary representation of the presence of any (A) maternal and (B) neonatal conditions.

For neonatal outcomes (**Fig. 3B**), there were 29 cases of small for gestational age (weight below the 10^th^ percentile; 16 Intervention, 13 Usual Care), 19 cases of large for gestational age (weight above the 90^th^ percentile; 9 Intervention, 10 Usual Care) and 30 cases of NICU admission (12 Intervention, 18 Usual Care). The incidence of NICU admission was lower in the Intervention Group (7% vs 11% in Usual Care; adjusted OR, 0.64; 95% CI, 0.29 to 1.38), which equated to an adjusted relative risk reduction of 28.6% (**eTable 4**).

### INTERVENTION ENGAGEMENT

Participants in the Intervention Group completed an average 16±4 check-ins with their coach. On average, 21±17 video lessons were viewed. Approximately two thirds of the participants engaged with self-monitoring behaviors on three or more days per week (**eTable 5**).

## DISCUSSION

The results of this trial show the effectiveness of a co-developed, fully remote prenatal lifestyle intervention for the attenuation of weight gain in pregnancy among low-income, predominantly Black pregnant individuals enrolled in WIC. Adherence to NAM guidelines was not different between Intervention and Usual Care Groups. However, the Intervention Group had a 13% average lower study observed total and rate of measured weight gain during pregnancy and a lower deviation from the NAM guidelines. Recommendations for optimal weight gain in pregnancy are actively debated because weight gain below current guidelines is often associated with the lowest comparative risk for adverse perinatal outcomes^2^. This is particularly emphasized in individuals with overweight and obesity. Indeed, a greater number of participants in the Intervention Group gained below NAM guidelines (23%) compared to Usual Care (18%).

The incidence of excess weight gain observed in the current trial are higher than those observed in epidemiological reports^12,13^. Compared to approximately 45%, 67% of our participants from low-income households gained more weight than recommendations, which underscores the exacerbated threat to pregnant people already facing the most serious risks for severe adverse maternal outcomes. Our effect size of 1.4 kg for total weight gain, achieved with a fully remote intervention, is consistent with the 1.0 kg effect size reported by the United States Preventive Services Taskforce (USPSTF)^14^. Of note, our findings differ from previous trials which tested fully remote interventions in WIC and observed no effect on gestational weight gain. The Healthy Beginnings Intervention – while fully remote – is considered high intensity (12 or more contacts) by the USPSTF^14^. Whereas these previous interventions in WIC utilized low intensity approaches, such as text message support^15^ or online self-directed resources^16^.

We hypothesized that the intervention would have equal impact for participants across all BMI categories. The pilot study for the present trial showed weight gain attenuation in individuals with obesity receiving the same multicomponent smartphone intervention but with intervention sessions via telehealth with a health coach^7^. With the fully remote intervention in the present trial, individuals with obesity did not experience the expected benefits as compared to those with normal weight. Obesity is a complex and multifactorial disease, and these data suggest remote interventions for this population require face-to-face support and delivery in primary care.

The NAM weight gain guidelines were established from five adverse perinatal outcomes including preterm birth^8^. Participants in the Intervention Group evidenced a clinically important reduction in the incidence of preterm birth compared to Usual Care. Preterm birth was experienced by 1 in 6 individuals in the Usual Care Group compared to 1 in 10 in the Intervention Group, equating to an adjusted relative risk reduction of 36.9%. A reduction in preterm birth incidence is a national priority. *Healthy People 2030* strive for a 1% reduction in preterm births by the year 2030^17^. A similar objective is claimed by the National WIC Association and WIC participation has been shown to reduce preterm births^18,19^. The National WIC Association is prioritizing an investment in technologies which would broaden its reach^6,20^. Thus, disseminating supportive weight management programs at scale could provide value added services historically beyond the scope of WIC and further reduce preterm births^19^.

Our trial has several strengths. Attrition not related to early delivery was low (16%), which we ascribe to three primary factors. First, the co-development of the intervention with our targeted population ensured relevance and a high level of engagement with each intervention component. Second, the fully remote format of the intervention allowed for continuity of care through the COVID-19 pandemic and four major hurricanes that impacted Louisiana and displaced participants. Third, as Black pregnant individuals in Louisiana experience the highest rates of maternal morbidity and mortality^21^, our statewide trial addresses the worsening disparities through inclusion of underserved communities notoriously absent in clinical research and most affected. Finally, as clinic assessment teams were deployed across the state, the trial was conducted rigorously without undue burden or interference with the WIC program.

Our trial also has limitations. The COVID-19 pandemic prohibited the trial from enrolling the a priori planned sample and equally across the nine geographic regions and BMI categories.

While the effect of pregnancy intervention trials on perinatal outcomes is critical to assess, sample sizes required for statistical inference are beyond the scope of such trials. The overall incidence of perinatal outcomes in the study sample align with state and national data which supports the clinical relevance of the trial findings^22^. Notably, the use of technology to remotely monitor weight and provide digital exercise and cooking videos was novel in 2019 when the trial launched. Accessibility to such resources expanded exponentially with COVID-19. Thus, it could be that digital resources available outside the trial influenced weight gain in both groups. Lastly, the intervention was delivered pragmatically, but the trial was not. Individuals having preexisting medical conditions including severe obesity or with barriers for trial completion were excluded.

## CONCLUSION

Remote delivery of a perinatal lifestyle intervention co-developed with low-income individuals receiving WIC supplemental nutrition significantly lowered gestational weight gain and reduced incidence of preterm birth.

Funding: This research was supported by funding from the National Institute of Nursing Research (5R01NR017644), the Louisiana/Pennington Nutrition and Obesity Research Center (NORC) of the National Institutes of Diabetes, Digestive and Kidney Diseases (P30DK072476) and the Louisiana Clinical and Translational Sciences Center (U54 GM104940).

## Data Availability

A deidentified dataset will be made publicly available in the Pennington/Louisiana NORCBiorepository at http://doi.org/10.17616/R31NJN8P

## Acknowledgements

The authors are indebted to the participants; support from the Louisiana Department of Health, and Louisiana Women, Infants and Children’s program staff; and health coaches and clinical assessment team, without whom this trial would not have been possible. The authors thank Dr. Kimberly Drews, the Director of Epidemiology and Research Design for the Louisiana Clinical and Translational Science Center, for her statistical oversight and guidance. Authors declare no conflicts of interest.

## Author Contributions

Redman had full access to all of the data in the study and takes responsibility for the integrity of the data and the accuracy of the data analysis.

Concept and design: Redman, Gilmore, Beyl.

Acquisition, analysis, or interpretation of data: All authors.

Drafting of the manuscript: Redman, Flanagan, Falkenhain.

Critical review of the manuscript for important intellectual content: All authors.

Statistical analysis: Beyl, Falkenhain.

Obtained funding: Redman.

Administrative, technical, or material support: Altazan.

Supervision: Redman, Apolzan, Gilmore, Altazan.

Other – specific input on Louisiana WIC mothers’ perspectives: Barlow.

