## Supplemental Appendix for "Gestational Weight Gain Management in Underserved Mothers - A State-Wide Randomized Controlled Trial in Louisiana WIC"

Supplementary Table S4a. Weight outcomes for participants with normal weight. ....................7

Supplementary Table S4b. Sensitivity Analysis - Weight outcomes for participants with normal weight and included weight data recorded at the mid-pregnancy outcome assessment for those with missing data at the end of pregnancy outcome assessment. ....................8

Supplementary Table S4c. Weight outcomes for participants with overweight. ....................9

Supplementary Table S4d. Sensitivity Analysis - Weight outcomes for participants with overweight and included weight data recorded at the mid-pregnancy outcome assessment for those with missing data at the end of pregnancy outcome assessment. ....................10

Supplementary Table S4e. Weight outcomes for participants with obesity. ....................11

Supplementary Table S4f. Sensitivity Analysis - Weight outcomes for participants with obesity and included weight data recorded at the mid-pregnancy outcome assessment for those with missing data at the end of pregnancy outcome assessment. ....................12

**Definition of Outcomes**

Per the 2009 guidelines of the National Academy of Medicine (NAM), each BMI category is allocated a different rate of gestational weight gain per week, which are the basis of our BMI-specific definitions (Table 1).

**Table 1. 2009 National Academy of Medicine Ranges for Weekly Gestational Weight Gain**

| BMI Category | Inadequate | Adequate | Excessive |
|---|---|---|---|
| **Normal Weight** | <0.35 kg/week | 0.35 kg/week to 0.50 kg/week | >0.50 kg/week |
| **Overweight** | <0.23 kg/week | 0.23 kg/week to 0.33 kg/week | >0.33 kg/week |
| **Obesity** | <0.17 kg/week | 0.17 kg/week to 0.27 kg/week | >0.27 kg/week |

Study-observed gestational weight gain *(secondary outcome)* is defined as the difference between clinic weight assessed during late pregnancy and clinic weight assessed during early pregnancy.

Weekly gestational weight gain *(secondary outcome)* is defined as study-observed gestational weight gain divided by the number of weeks between the early and late pregnancy study visits, with number of weeks calculated based on the number of days between study visits divided by 7.

Incidence of appropriate study-observed gestational weight gain per week *(primary outcome)* is defined as the number of participants that have weekly gestational weight gain in the adequate category according to the 2009 Institute of Medicine guidelines as opposed to inadequate or excessive (Table 1).

Deviation from the 2009 Institute of Medicine guidelines *(secondary outcome)* will be defined as the absolute difference in weekly gestational weight gain outside of the range bounds (Table 1). Participants within the bounds will not be included in this analysis.

Adverse perinatal outcomes *(exploratory outcomes)* were obtained from birth certificate records with the state. The Louisiana Electronic Event Registration System (LEERS) collects all official records related to birth and includes information from prenatal and delivery records.

Information abstracted from the LEERS records included the diagnosis of gestational diabetes, gestational hypertension/preeclampsia, and medically indicated Cesarean section as maternal outcomes. Neonatal intensive care unit admission, gestational age at delivery, and birth weight were used for calculation of small for gestational age or large for gestational age as neonatal outcomes. Specifically, medically indicated Cesarean section was defined as deliveries via Cesarean method of delivery with attempt of labor or if prior to 39 weeks with any medical indication. Small for gestational age (SGA, <10$^{th}$ percentile) and large for gestational age (LGA, >90$^{th}$ percentile) were defined using birth weight for gestations greater than 19 weeks and less than 45 weeks based on the Alexander criteria specific for neonatal sex and rate [4]. Preterm birth was defined as any birth occurring prior to 37 weeks gestation; the calculation of gestational age at delivery was determined as the difference in days between delivery date and the estimated date of conception divided by 7, with the delivery date abstracted from the birth certificate record and the date of conception calculated as 280 days prior to the participant self-reported due date.

From these outcomes, a maternal and neonatal composite was calculated. Specifically, for each participant, the number of adverse maternal outcomes (as listed above) was summed; from this, a binary maternal composite was derived with any participant who had a non-zero sum indicated as having a maternal adverse outcome and those with a zero-sum having no maternal adverse outcome. A neonatal composite was derived analogously based on the adverse neonatal outcomes (as listed above).

Other study measures (e.g., baseline characteristics) are briefly defined below:

Gestational age at randomization in weeks was defined as the number of days since the date of conception divided by 7, with the date of conception calculated as 280 days prior to the participant self-reported due date.

Parity: defined categorically as either nulliparous or non-nulliparous based on participant self-report.

Race/ethnicity: based on self-report and included the options 'Hispanic' (if participant reports any Hispanic, Latino/a, or Spanish origin), 'Non-Hispanic White' (if participant reports no Hispanic ethnicity and indicates White race), 'Non-Hispanic Black' (if participant reports no Hispanic ethnicity and indicates Black or African American race), and 'Mixed or Other' (if participant reports multiple races or ethnicities, or a race other than White or Black).

Marital status: defined based on self-report and included options 'Married/Living with Significant Other' (if participant reports to be married or living with significant other) or 'Not Married' (if participant reports not to be married, or to be separated, divorced, or widowed).

Educational attainment: defined based on self-report and included options 'Some college education' (if participant reports a college degree, postgraduate work, or 1-3 years of college, business, or technical school) or 'No college education' (if participant reports some high school or a high school diploma or GED).

**eTable 1. Baseline Characteristics of Study Participants With and Without Primary Outcome Data. ***

| Characteristic | Assessed (N=265) | Not Assessed (N=86) | P Value |
|---|---|---|---|
| **Gestational age at randomization – weeks** | 15.3±1.6 | 14.9±1.7 | 0.06 |
| **Maternal age – years** | 27±6 | 27±6 | 0.43 |
| **Nulliparous – no. (%)** | 115 (43) | 35 (41) | 0.75 |
| **Body mass – kg** | 75.1±16.6 | 76.6±17.6 | 0.47 |
| **Body mass index – kg/m² †** | 28.5 ± 5.6 | 29.3 ± 6.2 | 0.22 |
| Normal weight; 18.5-24.9 – no. (%) | 89 (34) | 25 (29) | 0.18 |
| Overweight; 25.0-29.9 – no. (%) | 79 (30) | 20 (23) | |
| Obesity; 30.0-40.0 – no. (%) | 97 (37) | 41 (48) | |
| **Race/Ethnicity – no. (%)** | | | |
| Hispanic | 21 (8) | 2 (2) | 0.06 |
| Non-Hispanic White | 85 (32) | 24 (28) | |
| Non-Hispanic Black | 143 (54) | 58 (67) | |
| Mixed or Other | 16 (6) | 2 (2) | |
| **Marital status – no. (%)** | | | |
| Married/Living with significant other | 150 (57) | 50 (58) | 0.90 |
| Not married | 115 (43) | 36 (42) | |
| **Educational attainment – no. (%)** | | | |
| Some college education | 171 (65) | 50 (58) | 0.35 |
| No college education | 94 (35) | 36 (42) | |

Baseline characteristics between participants with and without primary outcome data. *Plus-minus values are means ± SD. †The body-mass index is the weight in kilograms divided by the square of the height in meters.

**eTable 2a. Sensitivity Analysis - Weight outcomes included weight data recorded at the mid-pregnancy outcome assessment for those with missing data at the end of pregnancy outcome assessment.**

| | | | Intervention Effect | |
|---|---|---|---|---|
| | **Intervention (N=163)** | **Usual Care (N=154)** | **Adjusted Mean Difference (95% CI)** | **Adjusted Odds Ratio (95% CI)** |
| **Gestational weight gain per week – kg** § | 0.45 (0.42 to 0.49) | 0.51 (0.47 to 0.55) | -0.06 (-0.11 to -0.00) | |
| **Gestational weight gain deviation – kg** ‡ | 0.19 (0.16 to 0.22) | 0.24 (0.21 to 0.28) | -0.06 (-0.10 to -0.01) | |
| **Guideline attainment – no. (%)** ‡ | | | | |
| Recommended – no. (%) | 24 (17) | 21 (17) | | |
| Not recommended – no. (%) | 115 (83) | 105 (83) | | 1.00 (0.55 to 1.83) |
| Inadequate – no. (%) | 27 (23) | 19 (18) | | |
| Excess – no. (%) | 88 (77) | 86 (82) | | |

§Shown as the total weight change between early and last completed pregnancy outcome visit divided by the number of weeks between visits

‡Attainment and deviation of National Academy of Medicine (NAM) 2009 pregnancy weight guidelines

**eTable 2b. Sensitivity Analysis - Intervention engagement metrics included for participants with weight data recorded at the mid-pregnancy outcome assessment and missing data at the end of pregnancy outcome assessment. ***

| Engagement Metric | Low Engagement (<40%) | Moderate Engagement (40-70%) | High Engagement (>70%) |
|---|---|---|---|
| **Weighing** | | | |
| no. of participants (%) | 57 (35) | 55 (34) | 51 (31) |
| total no. of days across intervention | 25 ± 17 | 68 ± 18 | 101 ± 26 |
| average no. of days per week | 1.4 ± 0.9 | 3.8 ± 0.6 | 6.0 ± 0.7 |
| **Recording Steps** | | | |
| no. of participants (%) | 71 (44) | 25 (15) | 67 (41) |
| total no. of days across intervention | 13 ± 16 | 68 ± 25 | 115 ± 21 |
| average no. of days per week | 0.8 ± 0.9 | 4.0 ± 0.6 | 6.4 ± 0.6 |
| **Educational Videos** | | | |
| no. of participants (%) | 92 (57) | 47 (29) | 24 (15) |
| total no. of videos viewed | 7 ± 7 | 32 ± 5 | 43 ± 3 |
| **Coach contacts** | | | |
| no. of participants (%) | 8 (5) | 23 (14) | 132 (81) |
| total no. of coach contacts | 4 ± 3 | 10 ± 3 | 16 ± 4 |

*Plus-minus values are means ± SD. Shown are data from Intervention Group participants with mid- or late pregnancy outcome assessments available (N=163). Metrics are derived based on the first full intervention week following seven days after randomization, to account for shipping time of intervention toolkit, through the last complete week prior to the late pregnancy outcome assessment.

**eTable 3a. Weight outcomes for participants with normal weight.**

| | Intervention (N=47) | Usual Care (N=42) | Intervention Effect: Adjusted Mean Difference (95% CI) | Intervention Effect: Adjusted Odds Ratio (95% CI) |
|---|---|---|---|---|
| **Gestational weight gain – kg** † | 11.6 (10.1; 13.1) | 14.4 (12.8; 16.0) | -2.8 (-5.0; -0.6) | |
| **Gestational weight gain per week – kg** § | 0.55 (0.48; 0.62) | 0.67 (0.60; 0.75) | -0.12 (-0.22; -0.02) | |
| **Gestational weight gain deviation – kg**‡ | 0.13 (0.07; 0.19) | 0.28 (0.22; 0.35) | -0.15 (-0.24; -0.06) | |
| **Guideline attainment – no. (%)** ‡ | | | | |
| Recommended – no. (%) | 9 (19) | 10 (24) | | |
| Not recommended – no. (%) | 38 (81) | 32 (76) | | 0.76 (0.27 to 2.09) |
| Inadequate – no. (%) | 9 (24) | 4 (12) | | |
| Excess – no. (%) | 29 (76) | 28 (88) | | |

†Shown as the total weight change between early and late pregnancy assessment outcome visits

§Shown as the total weight change between early and late pregnancy assessment outcome visits divided by the number of weeks between visits

‡Attainment and deviation of National Academy of Medicine (NAM) 2009 pregnancy weight guidelines calculated from rate of weight gain per week.

**eTable 3b. Sensitivity Analysis - Weight outcomes for participants with normal weight and included weight data recorded at the mid-pregnancy outcome assessment for those with missing data at the end of pregnancy outcome assessment.**

| | | | Intervention Effect | |
|---|---|---|---|---|
| | Intervention (N=54) | Usual Care (N=52) | Adjusted Mean Difference (95% CI) | Adjusted Odds Ratio (95% CI) |
| **Gestational weight gain per week – kg§** | 0.56 (0.49; 0.62) | 0.66 (0.59; 0.72) | -0.10 (-0.19; -0.01) | |
| **Gestational weight gain deviation – kg‡** | 0.13 (0.08; 0.18) | 0.27 (0.21; 0.32) | -0.14 (-0.21; -0.06) | |
| **Guideline attainment – no. (%)** ‡ | | | | |
| Recommended – no. (%) | 9 (17) | 12 (23) | | 0.66 (0.25 to 1.74) |
| Not recommended – no. (%) | 45 (83) | 40 (77) | | |
| Inadequate – no. (%) | 9 (20) | 5 (12) | | |
| Excess – no. (%) | 36 (80) | 35 (88) | | |

§Shown as the total weight change between early and last completed pregnancy outcome visits divided by the number of weeks between visits

‡Attainment and deviation of National Academy of Medicine (NAM) 2009 pregnancy weight guidelines calculated from rate of weight gain per week.

**eTable 3c. Weight outcomes for participants with overweight.**

| | Intervention (N=42) | Usual Care (N=37) | Intervention Effect: Adjusted Mean Difference (95% CI) | Intervention Effect: Adjusted Odds Ratio (95% CI) |
|---|---|---|---|---|
| **Gestational weight gain – kg†** | 10.0 (8.4; 11.6) | 11.1 (9.4; 12.8) | -1.1 (-3.5; 1.2) | |
| **Gestational weight gain per week – kg** § | 0.46 (0.38; 0.53) | 0.53 (0.45; 0.60) | -0.07 (-0.18; 0.04) | |
| **Gestational weight gain deviation – kg** ‡ | 0.23 (0.17; 0.30) | 0.23 (0.17; 0.29) | 0.00 (-0.09; 0.09) | |
| **Guideline attainment – no. (%)** ‡ | | | | |
| Recommended – no. (%) | 9 (21) | 2 (5) | | |
| Not recommended – no. (%) | 33 (79) | 35 (95) | | 4.82 (0.97 to 24.10) |
| Inadequate – no. (%) | 5 (15) | 5 (14) | | |
| Excess – no. (%) | 28 (85) | 30 (86) | | |

†Shown as the total weight change between early and late pregnancy assessment outcome visits

§Shown as the total weight change between early and late pregnancy assessment outcome visits divided by the number of weeks between visits

‡Attainment and deviation of National Academy of Medicine (NAM) 2009 pregnancy weight guidelines calculated from rate of weight gain per week.

**eTable 3d. Sensitivity Analysis - Weight outcomes for participants with overweight and included weight data recorded at the mid-pregnancy outcome assessment for those with missing data at the end of pregnancy outcome assessment.**

| | | | Intervention Effect | |
|---|---|---|---|---|
| | Intervention (N=48) | Usual Care (N=41) | Adjusted Mean Difference (95% CI) | Adjusted Odds Ratio (95% CI) |
| **Gestational weight gain per week – kg §** | 0.45 [0.38; 0.52] | 0.49 [0.42; 0.57] | -0.04 [-0.14; 0.06] | |
| **Gestational weight gain deviation – kg ‡** | 0.22 [0.17; 0.28] | 0.23 [0.17; 0.28] | -0.00 [-0.08; 0.08] | |
| **Guideline attainment – no. (%) ‡** | | | | |
| Recommended – no. (%) | 9 (19) | 2 (5) | | |
| Not recommended – no. (%) | 39 (81) | 39 (95) | | 4.64 (0.94 to 23.00) |
| Inadequate – no. (%) | 7 (18) | 8 (21) | | |
| Excess – no. (%) | 32 (82) | 31 (79) | | |

§Shown as the total weight change between early and last completed pregnancy outcome visits divided by the number of weeks between visits

‡Attainment and deviation of National Academy of Medicine (NAM) 2009 pregnancy weight guidelines calculated from rate of weight gain per week.

**eTable 3e. Weight outcomes for participants with obesity.**

| | Intervention (N=50) | Usual Care (N=47) | Intervention Effect: Adjusted Mean Difference (95% CI) | Intervention Effect: Adjusted Odds Ratio (95% CI) |
|---|---|---|---|---|
| **Gestational weight gain – kg †** | 7.2 (5.7; 8.6) | 7.7 (6.2; 9.2) | -0.5 (-2.6; 1.6) | |
| **Gestational weight gain per week – kg §** | 0.36 (0.29; 0.43) | 0.38 (0.31; 0.45) | -0.02 (-0.12; 0.08) | |
| **Gestational weight gain deviation – kg ‡** | 0.21 (0.16; 0.27) | 0.23 (0.17; 0.29) | -0.02 (-0.10; 0.06) | |
| **Guideline attainment – no. (%) ‡** | | | | 0.58 (0.19 to 1.80) |
| Recommended – no. (%) | 6 (12) | 9 (19) | | |
| Not recommended – no. (%) | 44 (88) | 38 (81) | | |
| Inadequate – no. (%) | 13 (30) | 10 (26) | | |
| Excess – no. (%) | 31 (70) | 28 (74) | | |

†Shown as the total weight change between early and late pregnancy assessment outcome visits

§Shown as the total weight change between early and late pregnancy assessment outcome visits divided by the number of weeks between visits

‡Attainment and deviation of National Academy of Medicine (NAM) 2009 pregnancy weight guidelines calculated from rate of weight gain per week.

**eTable 3f. Sensitivity Analysis - Weight outcomes for participants with obesity and included weight data recorded at the mid-pregnancy outcome assessment for those with missing data at the end of pregnancy outcome assessment.**

| | | | Intervention Effect | |
|---|---|---|---|---|
| | Intervention (N=61) | Usual Care (N=61) | Adjusted Mean Difference (95% CI) | Adjusted Odds Ratio (95% CI) |
| **Gestational weight gain per week – kg** § | 0.35 (0.29; 0.41) | 0.38 (0.32; 0.44) | -0.03 (-0.12; 0.06) | |
| **Gestational weight gain deviation – kg** ‡ | 0.21 (0.16; 0.26) | 0.24 (0.19; 0.29) | -0.03 (-0.10; 0.04) | |
| **Guideline attainment – no. (%)** ‡ | | | | |
| Recommended – no. (%) | 8 (13) | 11 (18) | | |
| Not recommended – no. (%) | 53 (87) | 50 (82) | | 0.70 (0.26 to 1.90) |
| Inadequate – no. (%) | 16 (30) | 16 (32) | | |
| Excess – no. (%) | 37 (70) | 34 (68) | | |

§Shown as the total weight change between early and last completed pregnancy outcome visits divided by the number of weeks between visits

‡Attainment and deviation of National Academy of Medicine (NAM) 2009 pregnancy weight guidelines calculated from rate of weight gain per week.

**eTable 4. Incidence of perinatal outcomes for participants assigned to Intervention and Usual Care.**

| | Intervention (N=172) | Usual Care (N=171) | Adjusted Odds Ratio (95% CI) |
|---|---|---|---|
| **Maternal outcomes – no. (%)** | | | |
| Gestational diabetes | 13 (8) | 13 (8) | 1.04 (0.46 to 2.36) |
| Gestational hypertension | 26 (15) | 29 (17) | 0.80 (0.44 to 1.46) |
| Medically indicated Cesarean | 36 (21) | 37 (22) | 0.93 (0.54 to 1.59) |
| Preterm delivery (<37 weeks) | 16 (9) | 27 (16) | 0.56 (0.28 to 1.08) |
| Composite outcome | 68 (40) | 74 (43) | 0.82 (0.52 to 1.27) |
| **Fetal/Neonatal outcome – no. (%)** | | | |
| NICU admission | 12 (7) | 18 (11) | 0.64 (0.29 to 1.38) |
| Small for GA (<10%$^{th}$ percentile) | 16 (9) | 13 (8) | 1.14 (0.52 to 2.51) |
| Large for GA (>90%$^{th}$ percentile) | 9 (5) | 10 (6) | 0.92 (0.35 to 2.35) |
| Composite outcome | 35 (20) | 40 (23) | 0.81 (0.48 to 1.36) |

Shown are adjusted odds ratios. NICU denotes neonatal intensive care unit. GA denotes gestational age.

**eTable 5. Intervention Engagement Metrics. ***

| | Engagement <2 days per week | Engagement 3-5 days per week | Engagement >5 days per week |
|---|---|---|---|
| **Weighing** | | | |
| no. of participants (%) | 48 (35) | 48 (35) | 43 (31) |
| average no. of days per week | 1.5±0.9 | 3.8±0.5 | 5.9±0.7 |
| **Recording Steps** | | | |
| no. of participants (%) | 56 (40) | 20 (14) | 63 (45) |
| average no. of days per week | 0.8±0.9 | 4.1±0.6 | 6.4±0.6 |

*Plus-minus values are means ± SD. Shown are data from Intervention Group participants with late pregnancy outcome assessments available (N=139). Engagement for <2 days per week equates to less than 40% average weekly engagement; engagement for 3-5 days per week equates to 40-70% average weekly engagement; engagement for >5 days per week equates to at least 70% average weekly engagement.
